# Effect of iStent inject on unmedicated intraocular pressure in glaucoma: exploratory analysis of clinical trial data

**DOI:** 10.64898/2026.08.23.26361150

**Authors:** Zhengyang Liu, Jennifer C. Fan Gaskin, Ghee Soon Ang, Deus Bigirimana, George Yu Xiang Kong, Alp Atik, Myra B. McGuinness

**Author notes:** Corresponding Author* Myra B. McGuinness, Centre for Epidemiology and Biostatistics, Melbourne School of Population and Global Health, University of Melbourne. Level 3, 207 Bouverie St, Carlton, 3053, VIC Australia.

## Abstract

**Purpose:** The direct effect of iStent *inject* on intraocular pressure (IOP) in patients with glaucoma is difficult to quantify in pragmatic trials where rates of post-surgical IOP-lowering therapy differ between intervention groups. We aimed to quantify the causal effect of iStent *inject* on unmedicated IOP at 12- and 24-months post-surgery.

**Methods:** Adults with mild-to-moderate glaucoma were 1:1 randomised to receive cataract surgery with iStent *inject* or cataract surgery alone at an Australian hospital (2017-2020, NCT03106181). IOP-lowering medications were prescribed as per clinician discretion. An exploratory analysis was used to estimate the controlled direct effect of iStent *inject* on IOP, analogous to the effect expected if all participants had undergone medication washout prior to assessment.

**Results:** Ninety-five eyes from 80 people were included (67.4% male, mean age 73.0 years, mean baseline IOP 17.1 mmHg). IOP-lowering medication was required for 53% of eyes in each group at 12 months (n=76); at 24 months (n=86) it was required for 43% and 64% in the active and control groups, respectively. Mean IOP was similar between intervention groups at each outcome visit. The controlled direct effect favoured the iStent *inject* group at 12 months (-2.1-mmHg difference, 95% CI -4.0,-0.3) but was attenuated at 24 months (-0.5 mmHg-difference, 95% CI -2.6,1.6).

**Conclusion:** Although the iStent *inject* was estimated to have an effect on lowering unmedicated IOP at 12 months, this effect had largely disappeared by 24 months. Medication washout is recommended when safe and practical in future trials to estimate these direct effects with more certainty.

## Introduction

Intraocular pressure (IOP) reduction has been shown to slow disease progression and vision loss in glaucoma, and can be accomplished by various treatment modalities, including eye drops, laser therapy, and surgical interventions.^1^ The iStent *inject* Trabecular Micro-Bypass System (Glaukos Corporation, California, USA) was designed to implant two stents in the drainage angle of the eye to increase fluid outflow and reduce IOP.^2^ Used in combination with or without cataract surgery, this minimally invasive glaucoma surgery provides an alternative approach to glaucoma control, potentially offering comparable outcomes to traditional surgical interventions such as trabeculectomy.^3^

Early randomised studies found that iStent *inject* led to a sustained reduction in unmedicated IOP.^4,5^ A later pragmatic trial found no evidence of a difference in medicated IOP between intervention and control groups at the primary outcome visit, despite a reduced need for IOP-lowering medication in the iStent *inject* group compared to participants who underwent cataract surgery alone.^6^ Observational studies have similarly supported an IOP and topical medication reducing effect with iStent use.^7–13^ However, quantification of the effect of the iStent *inject* on IOP within pragmatic trials is complicated by the mediating effect of post-randomisation treatment with IOP lowering medication. In these trials where medication washout has not been imposed, excluding participants who require treatment escalation introduces bias by disrupting the intervention group balance achieved via randomisation, while including them may dilute the estimated treatment effect.^14^ Causal inference provides one approach to investigating the degree to which iStent *inject* impacts IOP through mechanisms other than those mediated by post-operative topical therapy (i.e., the “direct” effect of the intervention).^15^ The addendum to the ICH guideline on Statistical Principles for Clinical Trials (ICH E9 R1) describes treatment escalation as a type of “intercurrent event” and lists the “hypothetical strategy” as one approach that can be used in clinical trials to evaluate what the effect on an intervention would be in a setting where the intercurrent event could be prevented or imposed.^16,17^ Hence, we aimed to assess the effect of undergoing cataract surgery with iStent *inject* on IOP that is not mediated by the prescription of post-surgical IOP-lowering treatment using clinical trial data. That is, we estimated the mean difference in IOP at both 12 and 24 months between people with mild-moderate glaucoma who underwent cataract surgery combined with iStent *inject* and those who underwent cataract surgery alone in the hypothetical setting in which unmedicated IOP had been measured for all participants.

## Materials and methods

### Study design

The data were originally collected during a prospective, parallel group, assessor-masked controlled trial (clinicaltrials.gov ID NCT03106181).^6^ Patients attending the Glaucoma Research and Investigation Unit outpatient clinic at the Royal Victorian Eye and Ear Hospital (RVEEH) in Australia were screened for eligibility between 2017 and 2020.

Surgery was conducted at a single site (RVEEH) by multiple surgeons. The clinical trial protocol was approved by the RVEEH Human Research and Ethics Committee (17/1333H), and the study was conducted in compliance with the Declaration of Helsinki. Written informed consent was obtained from all participants prior to enrolment after the nature of the trial was explained. The trial had an initial recruitment target of 100 participants per group (200 total) to detect a 2-mmHg difference in IOP between groups. However, recruitment was stopped due to COVID-19 pandemic-related delays after 25 months when 111 eyes had been randomised. Findings relating to the primary, secondary, and safety objectives of the trial have been previously published.^6^ The current study involves exploratory post-hoc analyses.

### Eligibility criteria

In brief, adults with open angle glaucoma (including pigmentary and pseudoexfoliative glaucoma) and age-related cataract were eligible to be enrolled in the trial as previously described.^6^ Participants were required to have an IOP of 12 to 30 mmHg while on 0-3 IOP-lowering medications, and a Humphrey Field Analyzer 24-2 visual field mean deviation better than -16 dB at baseline. If both eyes were eligible for the study, then both eyes could be included and the eye with the poorer visual acuity was operated on first.

### Randomisation and intervention

Eyes were allocated 1:1 to either cataract surgery with iStent *inject* (active intervention) or cataract surgery alone (control) using software-generated simple randomisation without stratification. Participants and treating surgeons were aware of intervention group allocation prior to surgery.

All participants underwent phacoemulsification and intraocular lens insertion as per standard of care. Two stents (GTS-400, Glaukos Corporation, San Clemente, CA) were then implanted into Schlemm’s canal using the preloaded iStent *inject* Trabecular Micro-bypass System (G2-M-IS) for participants allocated to the active intervention arm. The first stent was implanted into the nasal angle and the second was implanted between two and three clock hours away. Three participants received three stents due to surgical difficulties and poor view.

Aqueous release was performed at one day post-op if the IOP was significantly elevated (n=3 in the control group) and was not included in the definition of treatment escalation for this study.

### Clinical assessment and treatment escalation

No medication washout was conducted prior to baseline or follow-up visits although IOP-lowering medications were stopped on the day of surgery. Postoperative study visits were planned for 1 day and at 1, 4, 10, 16, 22, 52, 78, and 104 weeks. For the current analysis, the 12- and 24-month data were taken from the visit that occurred closest to the target date (after the 22-week and before the 78-week visit for 12-months, after the 78-week visit for 24-months) and included visits that occurred outside the planned visit windows due to COVID-related disruptions.

“Treatment escalation” (i.e., prescription of IOP-lowering eye drops, oral acetazolamide [Diamox], laser, or surgery) was pragmatically permitted during the study period to simulate real-world conditions and was dichotomised for the current analysis as being prescribed by the outcome visit or not. The need for treatment escalation and the type, dose, frequency, and duration of IOP-lowering eye drop use were determined as per individual clinician judgement based on Goldmann applanation IOP, optic disc, and visual field findings (all completed while masked to intervention group prior to gonioscopy if it was performed) and may have been determined based on assessments conducted between study visits. Each active ingredient was counted as a separate IOP-lowering medication class (e.g., Cosopt [Mundipharma Pty Ltd] was recorded as two medication classes, dorzolamide and timolol). Data on compliance with IOP-lowering medications was not captured in the clinical trial data.

### Statistical analysis

The analysis set for each outcome visit included all participants who attended that visit. The distribution of each characteristic was summarised as mean (SD) for variables with an approximately normal distribution, median [IQR] for continuous variables with a non-normal distribution, and frequency (%) for categorical variables, and aggregated by intervention received (iStent *inject* or Control). Comparisons of baseline characteristics were made between intervention groups via two-sample t tests (for variables with an approximately normal distribution), Wilcoxon rank-sum test (continuous variables without a normal distribution), Fisher’s exact test (for categorical variables with any cell count <5), and Pearson’s chi-squared test (all other categorical variables).

A directed acyclic graph was used to depict assumptions about the causal relationships between the intervention (iStent *inject*), outcome (IOP), mediator (post-surgical treatment escalation), and potential mediator-outcome confounders, and to inform covariate selection for each of the models listed below (see Supplemental Figure 1).

Outcomes were assessed separately for 12- and 24-month visits. For each outcome visit, a weighted and baseline covariate-adjusted marginal structural model for IOP was fit with an interaction between surgical intervention group and treatment escalation status to estimate the controlled direct effect of iStent *inject* (i.e., the effect on unmedicated IOP that would be observed if washout had occurred for all participants).^18,19^ Stabilised inverse probability weights were derived to account for post-randomisation treatment escalation^18^ and to reduce the potential for bias due to missing data.^20^ To generate weights, the probability of requiring treatment escalation was estimated via logistic regression, adjusting for surgical group (iStent *inject* vs control), baseline characteristics (age, IOP, number of IOP-lowering medications, pseudoexfoliation status, and sex), and highest IOP prior to the outcome visit (a time-varying mediator-outcome confounder impacted by the surgical intervention).^21^ The probability of having non-missing data incorporated in the weights was estimated (among all operated participants for whom baseline data were collected) via logistic regression adjusting for surgical group and the baseline characteristics specified above. It is likely that the missing outcome data are “missing not at random” (as suggested by the directed acyclic graph in Supplemental Figure 1) with potential for residual bias.^22^ Weights were truncated at the 15^th^ and 85^th^ centiles to obtain a smaller range with mean closer to 1.0.^23^ As sensitivity analyses, the controlled direct effects were re-estimated, firstly with untruncated weights, and secondly without weighting for missing data. As a comparison, traditional unweighted linear regression was also used to estimate the baseline covariate-adjusted effect of iStent *inject* on IOP (i.e., the mean difference in follow-up IOP between intervention groups regardless of treatment escalation; the “treatment policy strategy” for handling intercurrent events^16^). Confidence intervals for all effect estimates were derived from robust standard errors, accounting for correlation between right and left eyes of participants who contributed both eyes to the analyses.

To facilitate comparisons with other studies and clinical guidelines,^24–26^ IOP at each follow-up visit was dichotomised according to whether it had reduced from baseline by at least 2 mmHg, 4 mmHg, or 20%.

Analyses were conducted using Stata/BE v19.0 (StataCorp, College Station, TX).

## Results

### Baseline characteristics

One hundred and eleven eyes from 93 participants were randomised, and 104 eyes from 87 participants underwent surgery. Two eyes randomised to the control group received the iStent *inject*. After excluding eyes that did not have either 12- or 24-month outcome data (n=9), 95 eyes from 80 people were included in one or both of the analysis sets (12 and 24 months, see flow chart, Supplemental Figure 2). Baseline characteristics were broadly similar between included and excluded participants, although baseline IOP was higher among excluded participants (Supplemental Table 1).

Forty-four of the included eyes underwent cataract surgery alone (46.3%) and 51 underwent cataract surgery with iStent *inject* (53.7%). Included participants were aged 53-84 years at time of surgery and 67.4% (n=64) were male. Baseline characteristics were similar between intervention groups, with mean (SD) baseline IOP of 16.9 mmHg (3.2) in the control group and 17.3 mmHg (3.8) in the active intervention group (see Table 1). Both groups had a median [IQR] of 2 [1,3] IOP-lowering medications (mean [SD] 1.8 [1.2] and 1.7 [1.1] in the control and active intervention groups, respectively).

**Table 1:** Baseline characteristics according to surgical intervention received.

|  | Cataract surgery<br>alone<br>(n=44) | Cataract surgery<br>& iStent *inject*<br>(n=51) | p-value* |
| --- | --- | --- | --- |
| Age (years), mean (SD) | 72.1 (7.6) | 73.8 (7.4) | 0.281 |
| IOP (mmHg), mean (SD) | 16.9 (3.2) | 17.3 (3.8) | 0.514 |
| Sex, n (%) |  |  |  |
| Female | 13 (29.5%) | 18 (35.3%) | 0.551 |
| Male | 31 (70.5%) | 33 (64.7%) |  |
| Race, n (%) |  |  |  |
| White | 31 (70.5%) | 40 (78.4%) | 0.721 |
| Asian | 9 (20.5%) | 8 (15.7%) |  |
| Other | 4 (9.1%) | 3 (5.9%) |  |
| Pseudoexfoliation, n (%) |  |  |  |
| Absent | 41 (93.2%) | 45 (88.2%) | 0.498 |
| Present | 3 (6.8%) | 6 (11.8%) |  |
| Number of medications, median [IQR] | 2.0 [1.0, 3.0] | 2.0 [1.0, 3.0] | 0.684 |
| Visual field mean deviation (dB), median [IQR] | -3.7 [-6.4, -2.2] | -4.1 [-8.7, -2.3] | 0.553 |
| Best corrected visual acuity (logMAR), median [IQR] | 0.18 (0.16) | 0.20 (0.20) | 0.707 |

\* p-values from two-sample t test (variables with approximately normal distribution), Wilcoxon rank-sum test (continuous variables without normal distribution), Fisher's exact test (race, pseudoexfoliation), or Pearson's chi-squared test (sex).

### Treatment escalation

Two of the participants in the iStent *inject* group were scheduled for trabeculectomy between their 12- and 24-month visits. Nine participants in the control group (20.5%) and six participants in the iStent *inject* group (11.7%) were given oral acetazolamide in the follow-up period. No IOP-lowering laser procedures were recorded (YAG posterior capsulotomy was indicated for one participant from each group at 12 months).

At 12 months the proportion of eyes requiring treatment escalation was similar between intervention groups (52.6% overall, see Table 2). At this time, the median number of medication classes [IQR] was 1 [0,2] in each intervention group. By 24 months, the proportion of eyes requiring IOP-lowering medication was greater in the control group (64.3%, median [IQR] number of classes 1.5 [0,3]) than in the iStent *inject* group (43.2%, 0 [0,1])

**Table 2:** Observed mean intraocular pressure, presented by surgical group and intraocular pressure-lowering treatment escalation status.

| Outcome visit | Intervention received | Treatment escalation | n (%) | Intraocular pressure (mmHg), Mean (SD) |  |  |  |
| --- | --- | --- | --- | --- | --- | --- | --- |
|  |  |  |  | Baseline | Interim* | Follow-up | Change |
| 12 months | Control | All | 36 | 17.0 (3.4) | 21.5 (7.2) | 15.2 (3.9) | -1.8 (3.4) |
|  |  | No | 17 (47.2) | 17.2 (3.8) | 19.6 (4.7) | 15.2 (2.8) | -2.0 (3.7) |
|  |  | Yes | 19 (52.8) | 16.9 (3.2) | 23.2 (8.6) | 15.2 (4.7) | -1.7 (3.4) |
|  | iStent *inject* | All | 40 | 17.4 (4.1) | 20.1 (8.4) | 14.5 (4.8) | -2.8 (4.3) |
|  |  | No | 19 (47.5) | 16.9 (3.3) | 18.4 (4.9) | 12.9 (2.8) | -4.0 (4.1) |
|  |  | Yes | 21 (52.5) | 17.7 (4.7) | 21.7 (10.6) | 16.0 (5.9) | -1.8 (4.3) |
|  | All | No | 36 (47.4) | 17.1 (3.5) | 19.0 (4.8) | 14.0 (3.0) | -3.0 (4.0) |
|  |  | Yes | 40 (52.6) | 17.3 (4.0) | 22.4 (9.6) | 15.6 (5.3) | -1.7 (3.9) |
| 24 months | Control | All | 42 | 17.0 (3.1) | 22.8 (7.1) | 14.9 (3.7) | -2.2 (3.6) |
|  |  | No | 15 (35.7) | 17.4 (3.4) | 20.3 (5.2) | 15.0 (3.3) | -2.4 (3.3) |
|  |  | Yes | 27 (64.3) | 16.9 (2.9) | 24.2 (7.6) | 14.8 (4.0) | -2.0 (3.8) |
|  | iStent *inject* | All | 44 | 16.7 (3.1) | 20.9 (7.6) | 15.2 (5.6) | -1.5 (6.1) |
|  |  | No | 25 (56.8) | 16.9 (3.3) | 20.4 (5.9) | 14.9 (4.4) | -2.1 (5.1) |
|  |  | Yes | 19 (43.2) | 16.4 (2.8) | 21.5 (9.6) | 15.7 (7.1) | -0.7 (7.4) |
|  | All | No | 40 (46.5) | 17.1 (3.3) | 20.4 (5.6) | 14.9 (4.0) | -2.2 (4.4) |
|  |  | Yes | 46 (53.5) | 16.7 (2.9) | 23.1 (8.5) | 15.2 (5.4) | -1.5 (5.5) |

n = number of eyes observed at each outcome visit.
\* Interim intraocular pressure is the highest value observed for each eye during follow-up prior to the outcome visit, and may have been measured with or without intraocular pressure-lowering medication.

The most common active ingredient at each outcome visit was timolol (in combination with ≥1 other medication in all but two eyes), followed by bimatoprost (see Supplemental Table 2).

### Intraocular pressure

On average, the IOP recorded at interim study visits was slightly higher in the control group than the iStent *inject* group (see Table 2). At 12 months, 27.8% of the control group and 52.5% of the iStent *inject* group had a reduction in IOP of ≥20% of their baseline level. At 24 months, the percentage of eyes with ≥20% reduction from baseline was more similar between groups (35.7% control and 43.2% iStent *inject*, see Supplemental Table 3). The mean IOP at 12- and 24-months was also similar between surgical intervention groups (Figure 1).

**Figure 1:**
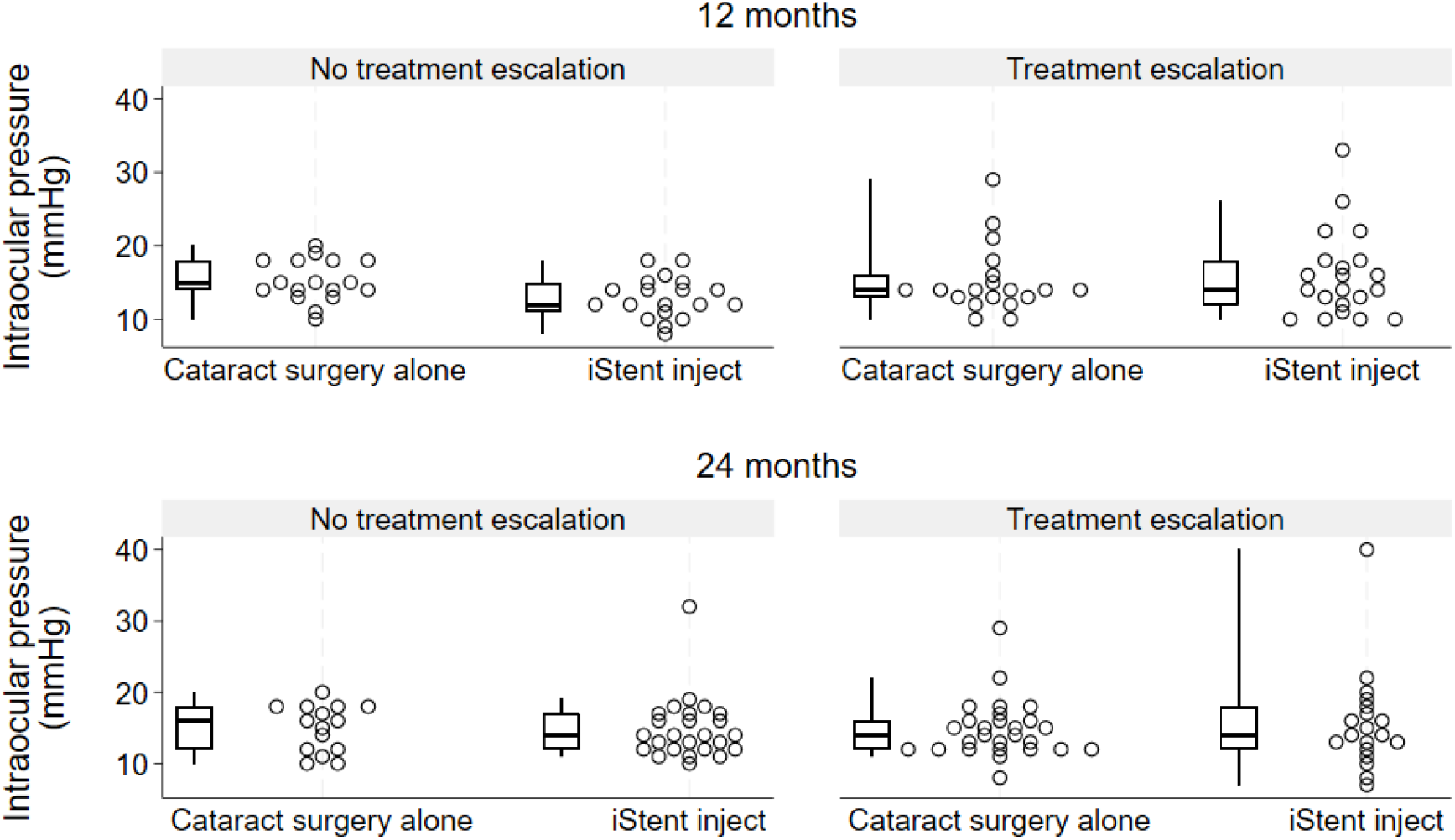
Distribution of intraocular pressure observed at outcome visits, stratified by surgical intervention and treatment escalation status. Each circle represents one eye. Boxes indicate median and interquartile range with whiskers extending to 5th and 95th centiles.

The adjusted mean difference in IOP between surgical intervention groups was -0.9 mmHg (95% CI -2.4, 0.6) at 12 months and 0.8 mmHg (95% CI -1.2, 2.9) at 24 months (i.e., insufficient evidence of an effect when ignoring treatment escalation).

The controlled direct effect of iStent *inject* at 12 months was estimated to be -2.2 mmHg (95% CI -4.0, -0.3), suggesting that if all participants had undergone medication washout, IOP would have been lower on average in the iStent *inject* group than the cataract surgery alone group. At 24 months there was less evidence of a difference between intervention groups (controlled direct effect -0.5 mmHg, 95% CI -2.6, 1.6). Results were similar when using alternative weighting schemes in the sensitivity analyses (see Supplemental Table 4).

## Discussion

In this randomised controlled trial of people with mild-to-moderate open angle glaucoma undergoing cataract surgery, a favourable effect of the iStent *inject* on unmedicated IOP was implied by the reduced need for IOP-lowering treatment at the primary outcome visit. Using causal inference methods, we found evidence of a direct effect of the iStent *inject* on unmedicated IOP at 12 months post-surgery, but were unable to confirm this at 24 months. The estimated 2.2 mmHg difference in unmedicated IOP between groups at 12 months is similar to values previously reported.^4,27^ It may be considered clinically relevant considering the measurement variability of Goldmann applanation tonometry,^28–30^ and the correlation between reduction in IOP and visual field sensitivity.^31^ The decrease in effect size between 12 and 24 months is consistent with previous assertions of decreased efficacy of minimally invasive glaucoma surgery over time.^32^

Guidelines suggest that a 20% reduction in IOP from baseline may be sufficient in the management of mild glaucoma.^25,26^ In the current study this was achieved without IOP-lowering medication for 12% of the cataract surgery alone group and 27% of the iStent *inject* group at 24 months; these proportions would likely be higher if pre-operative IOP had been assessed following medication washout.

We did not see a cumulative effect of combining the surgical and medical interventions in this study. The pharmacological pressure-lowering effects of eye drops may interact with the physiological mechanism of the stent, potentially reducing its impact on aqueous humour outflow, episcleral venous pressure, or other local physiological changes.^33–35^ Furthermore, patient variability, episcleral venous pressure, and compensatory mechanisms may alter IOP regulation when eye drops are used. These interactions underscore the complexity of glaucoma management, highlighting a need for further studies exploring how medical and surgical glaucoma therapies interact to affect long term IOP control.

Despite the cost of adding minimally invasive glaucoma surgery to phacoemulsification, health economic studies suggest that iStent insertion is cost-effective in a variety of healthcare systems.^36–39^ By decreasing the need for post-procedural treatment to achieve IOP control, the iStent improves patients’ quality of life. Future work is required to observe whether these gains are sustained over longer periods and the ultimate impact on functional measures of vision.

### Strengths and limitations

The strengths of this study include the random allocation of the surgical intervention, longitudinal data collection, and masked assessment of outcomes. Post-operative management was given as per standard of care so that results may be generalisable to real-world clinical practice.

A major limitation of this exploratory analysis is the sample size, with between 15 and 27 eyes in each stratum defined by surgical group and treatment escalation status. This limits the statistical power to decompose the interventional effect into both direct and indirect effects which are, by definition, smaller than the total average causal effect of the iStent *inject* on IOP.^40^ Although surgical intervention group was randomly allocated, an intention-to-treat approach was not followed due to participant attrition and inadvertent intervention switch. We aimed to reduce the potential impact of selection bias due to attrition using inverse probability weights, but residual bias may be present if people with lower final IOP were less likely to attend follow-up visits (i.e., if the outcome data are not missing at random). Furthermore, residual mediator-outcome confounding may have arisen from unmeasured or categorised baseline characteristics.

Valid causal inference requires certain assumptions to be met, including consistency of the mediator (i.e., that all participants with treatment escalation received the same dose of the medication).^41^ However, several different IOP-lowering agents were used and eye drop compliance was likely variable between participants.^42,43^ Thus, we were not able to reliably decompose the effect of iStent *inject* on IOP into direct and indirect effects. In addition, the positivity assumption (i.e., all eyes have non-zero probabilities of both receiving and not receiving treatment escalation) was likely violated given hospital protocols suggest prostaglandin analogues be commenced if IOP is ≥3 mmHg above baseline. Alternative analytic methods may be more suitable in these scenarios with deterministic treatment escalation.^17^

### Future research

Newer iStent delivery systems have been developed (iStent *inject* W, iStent infinite) and results may differ between models.

In future, the use of large observational datasets or individual participant data meta-analyses may provide sufficient power to explore this research question in more depth. As recommended for pre-market studies of minimally invasive glaucoma devices,^24^ the feasibility of medication washout prior to IOP assessment should be considered when designing future clinical trials.^32^ This would allow the direct effect of surgical interventions to be estimated with more certainty, and without the need for sophisticated statistical methodology or assumptions. However, it is acknowledged that medication washout may not be safe in trials of people with more severe disease, and that the primary objective of some trials may be to assess the reduction in medication burden.

## Conclusion

As previously reported, combined iStent *inject* surgery resulted in lower rates of treatment escalation compared to cataract surgery alone at the 24-month visit. Although a direct effect of the iStent *inject* on IOP was observed at 12 months, this had largely disappeared by the 24-month visit. We recommend IOP be measured following washout when practical in future trials to estimate these direct effects with more certainty.

## Data Availability

Data produced in the present study are available upon reasonable request to the authors

## Author contributions

Conception and design of clinical trial: JFG, GSA, GYXK. Data acquisition: JFG, DB, GYXK, AA, GSA. Analysis of data: ZL, MBM. ZL created the first draft of the paper. All authors contributed to interpretation of the data, reviewed the draft critically for intellectual content, gave final approval of the version to be published, and agree to be accountable for the work.

