## Supplemental Figure for "Effect of iStent inject on unmedicated intraocular pressure in glaucoma: exploratory analysis of clinical trial data"

### Online Supplement

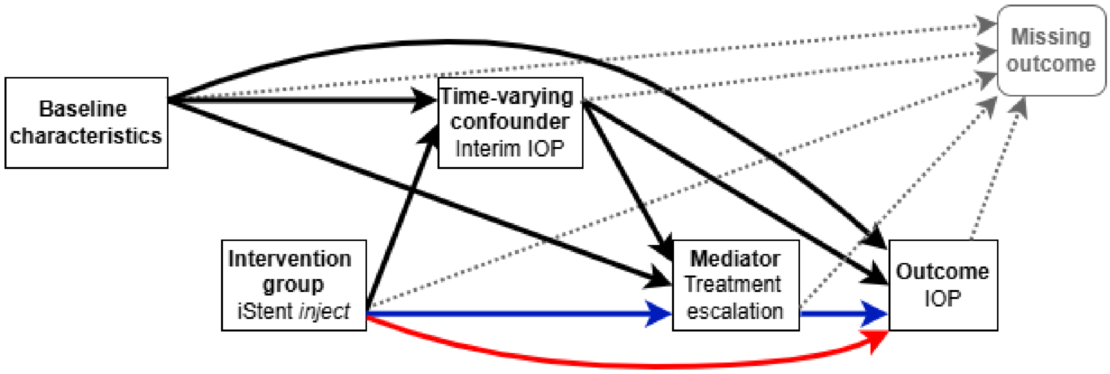


Supplemental Figure 1: Directed acyclic graph for the relationship between iStent *inject* and intraocular pressure (IOP) at 12 or 24 months. The direct effect pathway goes straight from intervention group to outcome (red arrow), while the indirect pathway travels from intervention group to outcome via the mediator (blue arrows). Characteristics with potential to affect both treatment escalation status and IOP were considered mediator-outcome confounders (age, IOP, number of IOP-lowering medications, pseudoexfoliation status, sex, intervention group, and highest IOP at interim visits). Baseline characteristics that may have an effect on treatment escalation status but not IOP (e.g., visual field and optical coherence tomography parameters) were not included in analysis models.

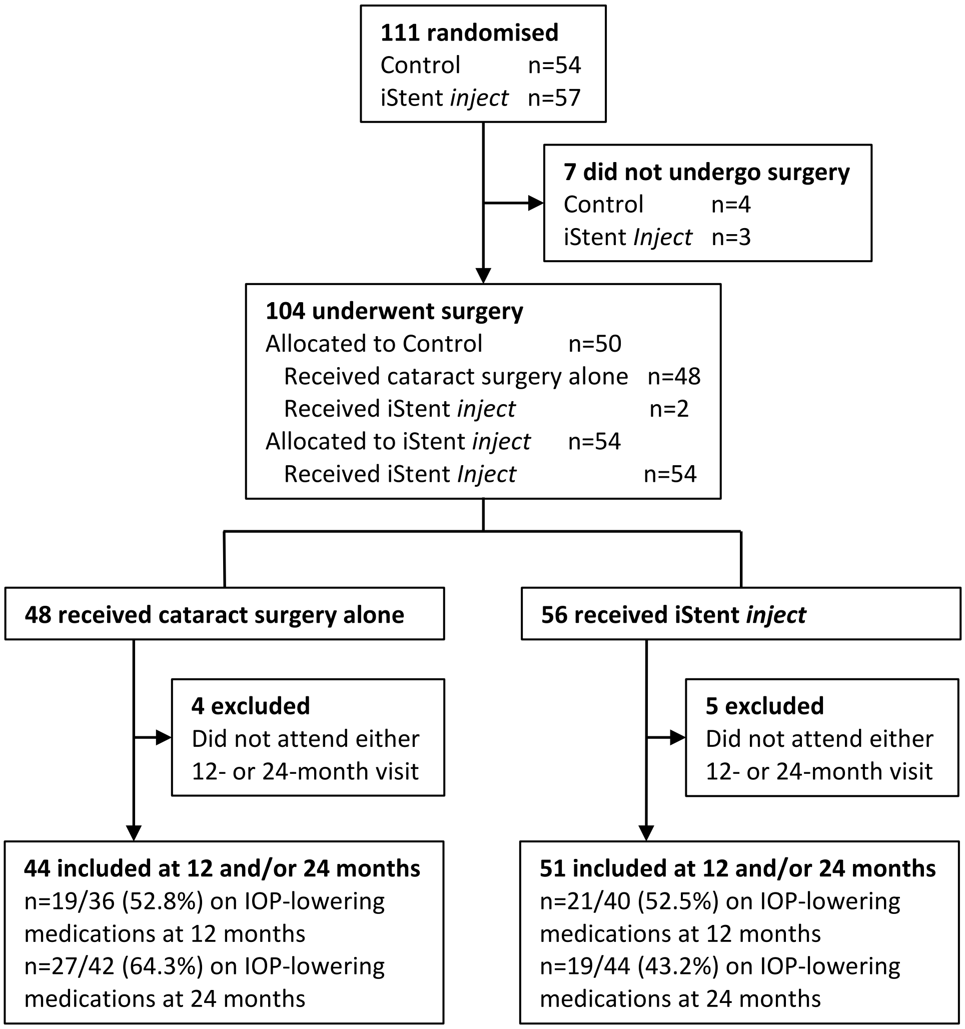


Supplemental Figure 2: Study flow diagram. IOP = intraocular pressure. Control group = cataract surgery alone; iStent *inject* = cataract surgery combined with iStent *inject*.

Supplemental Table 1: Baseline characteristics of excluded participants

| | 12 months | | | 24 months | | |
|---|---|---|---|---|---|---|
| | **Excluded** (n=28) | **Included** (n=76) | **p*** | **Excluded** (n=18) | **Included** (n=86) | **p*** |
| Study intervention received | | | | | | |
| Cataract surgery alone | 12 (42.9%) | 36 (47.4%) | 0.825 | 6 (33.3%) | 42 (48.8%) | 0.301 |
| Cataract surgery & iStent *inject* | 16 (57.1%) | 40 (52.6%) | | 12 (66.7%) | 44 (51.2%) | |
| Age (years) | 73.0 (6.8) | 73.3 (7.7) | 0.878 | 73.1 (8.2) | 73.3 (7.3) | 0.918 |
| Intraocular pressure (mmHg) | 18.3 (3.6) | 17.2 (3.7) | 0.200 | 20.5 (5.0) | 16.9 (3.1) | <0.001 |
| Sex | | | | | | |
| Female | 6 (21.4%) | 27 (35.5%) | 0.236 | 2 (11.1%) | 31 (36.0%) | 0.051 |
| Male | 22 (78.6%) | 49 (64.5%) | | 16 (88.9%) | 55 (64.0%) | |
| Self-identified race | | | | | | |
| White | 22 (78.6%) | 56 (73.7%) | 0.467 | 13 (72.2%) | 65 (75.6%) | 0.738 |
| Asian | 3 (10.7%) | 15 (19.7%) | | 3 (16.7%) | 15 (17.4%) | |
| Other | 3 (10.7%) | 5 (6.6%) | | 2 (11.1%) | 6 (7.0%) | |
| Pseudoexfoliation | | | | | | |
| Absent | 23 (82.1%) | 71 (93.4%) | 0.128 | 16 (88.9%) | 78 (90.7%) | 0.683 |
| Present | 5 (17.9%) | 5 (6.6%) | | 2 (11.1%) | 8 (9.3%) | |
| Number of medications | 2.0 [1.0, 2.0] | 2.0 [1.0, 3.0] | 0.679 | 2.0 [1.0, 2.0] | 2.0 [1.0, 3.0] | 0.712 |
| Central corneal thickness (microns) | 553.9 (36.9) | 534.8 (36.7) | 0.022 | 547.4 (35.7) | 538.5 (38.0) | 0.364 |
| Visual field mean deviation (dB) | -2.9 [-6.3, -1.5] | -4.5 [-8.5, -2.3] | 0.162 | -3.8 [-9.3, -0.9] | -3.8 [-6.8, -2.2] | 0.853 |
| Best corrected visual acuity (logMAR) | 0.2 (0.1) | 0.2 (0.2) | 0.789 | 0.3 (0.2) | 0.2 (0.2) | 0.144 |

Values are frequency (%), mean (SD), or median [IQR].
* p-values from two-sample t test (variables with approximately normal distribution), Wilcoxon rank-sum test (continuous variables without normal distribution), or Fisher's exact test (categorical variables). Central corneal thickness missing for 4 included eyes.

Supplemental Table 2: Intraocular pressure lowering medication at follow-up visits, by intervention received.

| | 12 months | | | 24 months | | |
|---|---|---|---|---|---|---|
| | **Control** (n=36) | **iStent *inject*** (n=40) | **Total** (n=76) | **Control** (n=42) | **iStent *inject*** (n=44) | **Total** (n=86) |
| Acetazolamide | 1 (2.8%) | 0 (0.0%) | 1 (1.3%) | 0 (0.0%) | 0 (0.0%) | 0 (0.0%) |
| Bimatoprost | 7 (19.4%) | 8 (20.0%) | 15 (19.7%) | 10 (23.8%) | 6 (13.6%) | 16 (18.6%) |
| Brimonidine | 3 (8.3%) | 8 (20.0%) | 11 (14.5%) | 9 (21.4%) | 3 (6.8%) | 12 (14.0%) |
| Brinzolamide | 6 (16.7%) | 5 (12.5%) | 11 (14.5%) | 6 (14.3%) | 2 (4.5%) | 8 (9.3%) |
| Dorzolamide | 0 (0.0%) | 2 (5.0%) | 2 (2.6%) | 3 (7.1%) | 2 (4.5%) | 5 (5.8%) |
| Latanoprost | 5 (13.9%) | 5 (12.5%) | 10 (13.2%) | 6 (14.3%) | 6 (13.6%) | 12 (14.0%) |
| Pilocarpine | 0 (0.0%) | 0 (0.0%) | 0 (0.0%) | 1 (2.4%) | 0 (0.0%) | 1 (1.2%) |
| Timolol | 13 (36.1%) | 13 (32.5%) | 26 (34.2%) | 18 (42.9%) | 7 (15.9%) | 25 (29.1%) |
| Travoprost | 6 (16.7%) | 6 (15.0%) | 12 (15.8%) | 7 (16.7%) | 4 (9.1%) | 11 (12.8%) |

Control = cataract surgery alone; iStent inject = cataract surgery and iStent inject.
Each eye had between 0 and 4 intraocular pressure lowering medications.

Supplemental Table 3: Proportion reaching thresholds for reduction in intraocular pressure from baseline.

| **Outcome visit** | **Intervention received** | **Treatment escalation** | **N** | **Reduction from baseline**, n (%) | | |
|---|---|---|---|---|---|---|
| | | | | **2 mmHg** | **4 mmHg** | **20%** |
| 12 months | Control | **All** | **36** | **22 (61.1)** | **9 (25.0)** | **10 (27.8)** |
| | | No | 17 | 10 (58.8) | 5 (29.4) | 5 (29.4) |
| | | Yes | 19 | 12 (63.2) | 4 (21.1) | 5 (26.3) |
| | iStent *inject* | **All** | **40** | **26 (65.0)** | **18 (45.0)** | **21 (52.5)** |
| | | No | 19 | 14 (73.7) | 11 (57.9) | 12 (63.2) |
| | | Yes | 21 | 12 (57.1) | 7 (33.3) | 9 (42.9) |
| | All | No | 36 | 24 (66.7) | 16 (44.4) | 17 (47.2) |
| | | Yes | 40 | 24 (60.0) | 11 (27.5) | 14 (35.0) |
| 24 months | Control | **All** | **42** | **19 (45.2)** | **13 (31.0)** | **15 (35.7)** |
| | | No | 15 | 6 (40.0) | 5 (33.3) | 5 (33.3) |
| | | Yes | 27 | 13 (48.1) | 8 (29.6) | 10 (37.0) |
| | iStent *inject* | **All** | **44** | **28 (63.6)** | **17 (38.6)** | **19 (43.2)** |
| | | No | 25 | 16 (64.0) | 10 (40.0) | 12 (48.0) |
| | | Yes | 19 | 12 (63.2) | 7 (36.8) | 7 (36.8) |
| | All | No | 40 | 22 (55.0) | 15 (37.5) | 17 (42.5) |
| | | Yes | 46 | 25 (54.3) | 15 (32.6) | 17 (37.0) |

N = number of eyes observed at each outcome visit, n (%) is the number (percent) within that group who achieved each level or greater reduction in intraocular pressure from baseline.

Supplemental Table 4: Sensitivity analyses.

| **Estimand*** | **Outcome visit,** *months* | **n** | **Inverse probability weighting** | | | **Mean difference (95% CI)** |
|---|---|---|---|---|---|---|
| | | | **Treatment escalation** | **Non-missing outcome** | **Truncation** | |
| Controlled direct effect | 12 | 76 | Yes | Yes | Yes | -2.2 (-4.0,-0.3) |
| | | | Yes | Yes | No | -2.1 (-3.9,-0.4) |
| | | | Yes | No | No | -2.1 (-3.8,-0.4) |
| | 24 | 86 | Yes | Yes | Yes | -0.5 (-2.6,1.6) |
| | | | Yes | Yes | No | -0.5 (-2.5,1.4) |
| | | | Yes | No | No | -0.5 (-2.4,1.4) |
| Baseline covariate-adjusted mean difference | 12 | 76 | No | No | No | -0.9 (-2.4,0.6) |
| | 24 | 86 | No | No | No | 0.8 (-1.2,2.9) |

Negative mean difference values indicate lower intraocular pressure (mmHg) in the iStent *inject* group than the control (cataract surgery alone) group.
n = number of eyes observed at each outcome visit.
* The controlled direct effect utilises the "hypothetical" strategy for dealing with the intercurrent event of treatment escalation, and represents the difference between groups if no participant was on intraocular pressure-lowering medication. The adjusted mean difference is estimated using the "treatment policy" strategy, where groups are compared regardless of the occurrence of the intercurrent event.
